# Circulating tumor DNA dynamics using a standardized multi-gene panel in advanced breast cancer treated with CDK4/6 inhibition and endocrine therapy

**DOI:** 10.1101/2020.06.29.20142257

**Authors:** Olga Martínez-Sáez, Tomás Pascual, Fara Brasó-Maristany, Nuria Chic, Blanca González-Farré, Esther Sanfeliu, Adela Rodríguez, Débora Martínez, Patricia Galván, Anna Belén Rodríguez, Francesco Schettini, Benedetta Conte, Maria Vidal, Barbara Adamo, Antoni Martinez, Montserrat Muñoz, Reinaldo Moreno, Patricia Villagrasa, Fernando Salvador, Eva M. Ciruelos, Iris Faull, Justin I. Odegaard, Aleix Prat

**Affiliations:** SOLTI Breast Cancer Research Group, Barcelona, Spain; Department of Medical Oncology, Hospital Clinic of Barcelona, Barcelona, Spain; Translational Genomics and Targeted Therapeutics in Solid Tumors, August Pi i Sunyer Biomedical Research Institute (IDIBAPS), Barcelona, Spain; Lineberger Comprehensive Cancer Center, University of North Carolina at Chapel Hill, Chapel Hill, NC, USA; Department of Pathology, Hospital Clinic of Barcelona, Barcelona, Spain; Department of Clinical Medicine and Surgery, University of Naples Federico II, Naples, Italy; Department of Medical Oncology U·O. Oncologia Medica 2, IRCCS Ospedale Policlinico San Martino, Largo R. Benzi 10, 16132, Genova, Italy; Department of Medical Oncology, Hospital 12 de Octubre, Madrid, Spain; Guardant Health, Inc., Redwood City, California

## Abstract

Circulating tumor DNA (ctDNA) levels may predict response to anticancer drugs^1^, including CDK4/6 inhibitors and endocrine therapy combinations (CDK4/6i+ET)^2^; however, critical questions remain unanswered such as which assay or statistical method to use^3^. Here, we obtained paired plasma samples at baseline and week 4 in 45 consecutive patients with advanced breast cancer treated with CDK4/6i+ET. ctDNA was detected in 96% of cases using the 74-gene Guardant360 assay^4,5^. A variant allele fraction ratio (VAFR) was calculated for each of the 79 detected mutations between both time-points. Mean of all VAFRs (mVAFR) was computed for each patient. In our dataset, mVAFR was significantly associated with progression-free survival (PFS). Baseline VAF, on-treatment VAF or absolute changes in VAF were not associated with PFS. These findings demonstrate that ctDNA dynamics using a standardized multi-gene panel and a unique methodological approach predicts treatment outcome. Clinical trials in patients with an unfavorable ctDNA response are needed.

---

In hormone receptor-positive (HR+) and HER2-negative advanced breast cancer, CDK4/6i+ET has remarkably improved survival outcomes and is now considered a standard treatment for most patients^6-8^. Although this is good news for patients suffering from metastatic breast cancer, improving the efficacy of CDK4/6i+ET using novel treatment strategies might be challenging. On one hand, no predictive biomarker exists to date to select patients who are going to progress early (i.e. the first 12 months) following CDK4/6i+ET^9^. On the other hand, improving survival outcomes with new or additional therapies when the control arm has a median PFS of 25-27 months in the first-line setting^10-12^ will require huge personal, physical and economic resources as well as long periods. This issue is not restricted to advanced breast cancer but also other cancer-types such as lung cancer.

Detection of ctDNA levels before and during therapy might improve CDK4/6i+ET efficacy, stratify patients and help design future trials with novel treatment strategies^3^. O’Leary and colleagues^2^ evaluated early ctDNA dynamics in patients with *PIK3CA*-mutated HR+/HER2-negative metastatic breast cancer treated with palbociclib and fulvestrant in PALOMA-3 trial. A multiplex digital PCR assay was used and hotspot *PIK3CA* mutations in exons 9 and 20 were evaluated in plasma^2^. *PIK3CA* mutations levels from baseline to day 15 of therapy were associated with PFS independently of the treatment received^2^. However, only 22% of patients with HR+/HER2-negative advanced breast cancer had detectable *PIK3CA* mutations in plasma. To circumvent this problem, others argue that individualized gene panels according to each patient’s tumor’s genetic profile should be prioritized^13^. However, single gene- or multiple assay-based approaches have important limitations and challenges for drug development, clinical trial design and clinical implementation.

We hypothesized that a standardized plasma-based sequencing assay that analyzes multiple genes simultaneously at baseline and after 4 weeks (i.e. cycle 2 day 1 [C2D1]) of CDK4/6i+ET can identify patients with HR+/HER2-negative advanced disease with different treatment outcomes. To accomplish this, we undertook a prospective study from May 2016 to June 2019 of 50 consecutive pre and postmenopausal patients with metastatic HR+/HER2-negative breast cancer treated as per standard practice with ET in combination with a CDK4/6 inhibitor (i.e. palbociclib or ribociclib) (**Fig. 1A**). Plasma samples were sequenced using the standardized Guardant360 assay v2.11 (Guardant Health, Inc., Redwood City, California)^5^, which can identify single nucleotide variants (SNVs) and indels from 74-genes (**Fig. 1B**). Among 50 patients, 2 patients (4%) had insufficient plasma volume, 2 patients (4%) had missing samples and 1 patient (2%) was treated in the adjuvant setting after resection of a supraclavicular lymph node and was excluded. Finally, 45 patients (90%) were evaluable of whom 31 (69%) had ctDNA-positive disease (i.e. highest VAF detected ≥VAF 0.4%) and 14 (31%) had ctDNA-low disease (i.e. highest VAF detected below 0.4%) at baseline (**Fig. 1C** and **Table 1**). Mutations in 42 genes were identified at baseline (27 with ≥VAF 0.4%) and the 4 most frequent altered genes were *PIK3CA, ESR1, TP53* and *ATM* (**Fig. 1D**); ≥1 mutation with ≥VAF 0.4% in any of these 4 genes was found in 24 patients (53.3%). As shown in previous studies^14,15^, all intrinsic molecular subtypes were identified in 27 HR+/HER2-negative tumor samples using the PAM50 subtype predictor^16^, although Luminal A and B subtypes predominated (**Fig. 1E**).

**Table 1.** Clinical features of the patient dataset according to ctDNA levels and dynamics.

|  | All patients<br>N = 45 | ctDNA-low<br>N = 14 | mVAFR-low<br>N = 11 | mVAFR-med/high<br>N = 20 |
| --- | --- | --- | --- | --- |
| **Age** |  |  |  |  |
| Median (range) yr | 61.4 (39-87) | 59.7 (39-87) | 56.9 (45-72) | 64 (42-75) |
| < 65 yr – no. (%) | 28 (62%) | 8 (57%) | 9 (82%) | 11 (55%) |
| ≥ 65 yr – no. (%) | 17 (38%) | 6 (43%) | 2 (18%) | 9 (45%) |
| **Line – no. (%)** |  |  |  |  |
| First | 21 (47%) | 8 (57%) | 5 (45%) | 9 (45%) |
| Second | 16 (36%) | 5 (36%) | 5 (45%) | 5 (25%) |
| Third or more | 8 (18%) | 1 (7%) | 1 (9%) | 6 (30%) |
| **ECOG-PS – no. (%)** |  |  |  |  |
| 0 | 17 (38%) | 5 (36%) | 5 (45%) | 7 (35%) |
| 1 | 27 (60%) | 9 (64%) | 6 (55%) | 12 (60%) |
| 2 | 1 (2%) | 0 | 0 | 1 (5%) |
| **Endocrine therapy – no. (%)** |  |  |  |  |
| Aromatase inhibitor | 15 (33%) | 4 (29%) | 3 (27%) | 8 (40%) |
| Fulvestrant | 26 (58%) | 9 (64%) | 7 (64%) | 10 (50%) |
| Tamoxifen | 4 (9%) | 1 (7%) | 1 (9%) | 1 (5%) |
| **Type of CDK4/6 inhibitor – no. (%)** |  |  |  |  |
| Palbociclib | 40 (89%) | 12 (86%) | 10 (91%) | 18 (90%) |
| Ribociclib | 5 (11%) | 2 (14%) | 1 (9%) | 2 (10%) |
| **Disease site – no. (%)** |  |  |  |  |
| Visceral | 29 (64%) | 10 (71%) | 7 (64%) | 12 (60%) |
| Non visceral | 16 (36%) | 4 (29%) | 4 (36%) | 8 (40%) |
| Bone-only | 10 (22%) | 1 (7%) | 4 (36%) | 5 (25%) |
| **Number of metastatic locations – no. (%)** |  |  |  |  |
| <3 | 17 (38%) | 4 (29%) | 4 (36%) | 7 (35%) |
| ≥3 | 28 (62%) | 10 (71%) | 7 (64%) | 13 (65%) |
| **“De novo” metastasis – no. (%)** |  |  |  |  |
| No | 33 (73%) | 10 (71%) | 7 (64%) | 15 (75%) |
| Yes | 12 (27%) | 4 (29%) | 4 (36%) | 5 (25%) |
| **Prior hormone sensitivity – no. (%)** |  |  |  |  |
| Sensitivity | 31 (69%) | 9 (64%) | 7 (64%) | 15 (75%) |
| Resistance* | 14 (31%) | 5 (36%) | 4 (36%) | 5 (25%) |

PS: performance status
\*Hormone resistance defined as relapse while on the first 2 years of adjuvant ET, or progression disease within first 6 months of first-line ET for advanced breast cancer, while on ET.

**Fig 1.**
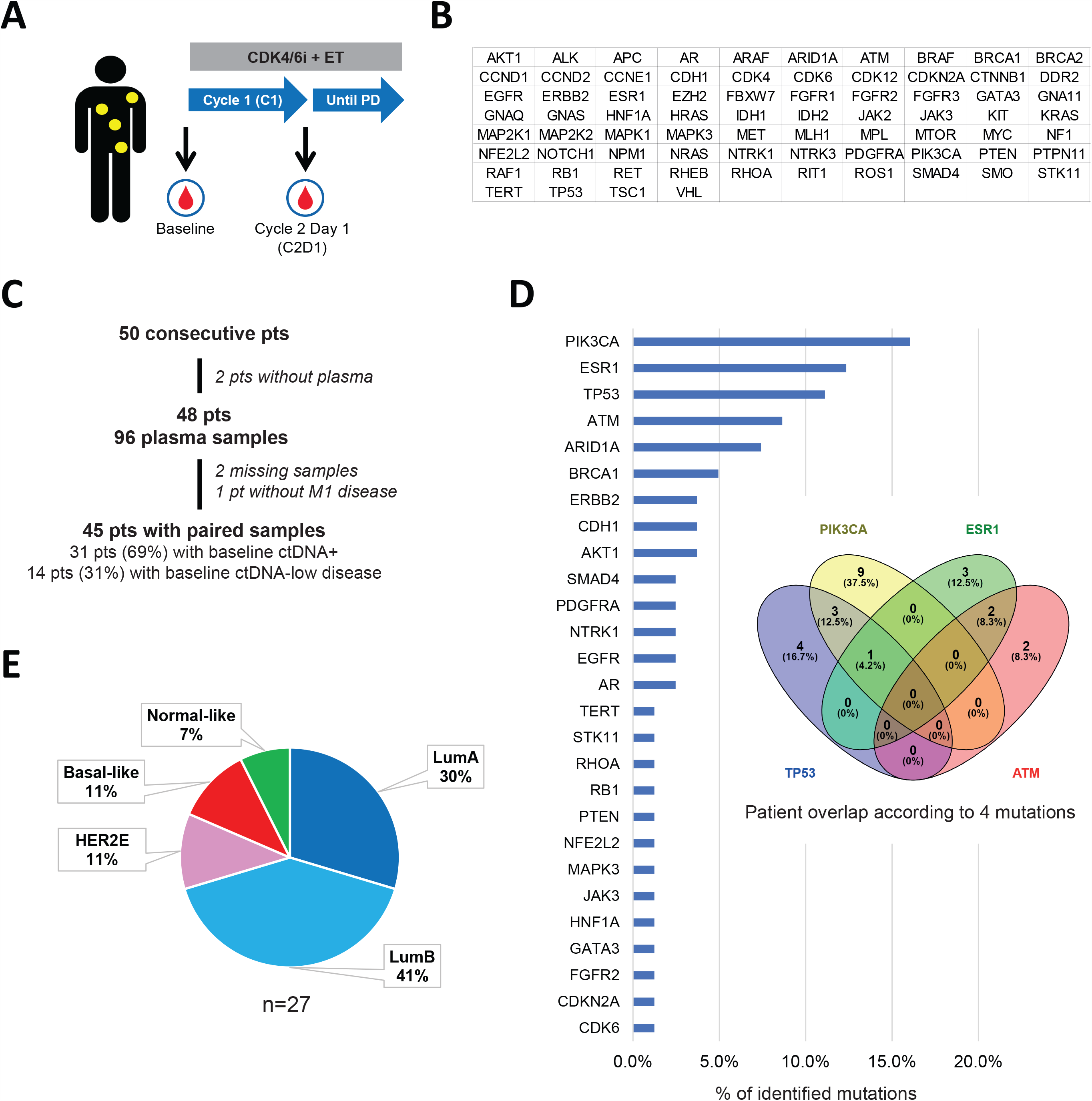
Description of the study. **(A)** Blood samples were extracted at baseline and after 1 cycle (i.e. 4 weeks) in patients with metastatic HR+/HER2-negative breast cancer treated with CDK4/6+ET; **(B)** The list of 74-genes analyzed by Guardant360; **(C)** CONSORT diagram; **(D)** Frequency of gene mutations with ≥VAF 0.4% at baseline identified in the patient dataset and Venn diagram with the 4 most frequent mutations; **(E)** PAM50 distribution (LumA=Luminal A, LumB=Luminal B, HER2E=HER2-Enriched).

A total of 79 single genetic mutations (VAF range 0.4-46%) were found at baseline in 31 patients (mean of 2.6 alterations per patient). All single mutations were tracked at C2D1. Mean VAF (mVAF) of the 79 mutations was 6.2 at baseline and 5.1 at C2D1 (p-value=0.040) (**Fig. 2A**). Any decrease in VAF at C2D1 compared to baseline was observed in 71% (56/79) of the tracked mutations. To capture the magnitude of ctDNA response, a VAFR from C2D1 to baseline was calculated for each genetic mutation and a mVAFR was computed for each patient (**Fig. 2B**). The range of mVAFR was 0.1 to 4.5 and 34% of patients had mVAFR of ≤0.3 (mVAFR-low), 28% had mVAFR of 0.31-0.99 (mVAFR-medium) and 38% had a mVAFR of ≥1.0 (mVAFR-high) (**Fig. 2C**). Of note, 1 patient (7.1%) with ctDNA-low disease at baseline had ctDNA-positive disease at C2D1. Finally, no clinical features were found specific of a particular ctDNA group (**Table 1**).

**Fig 2.**
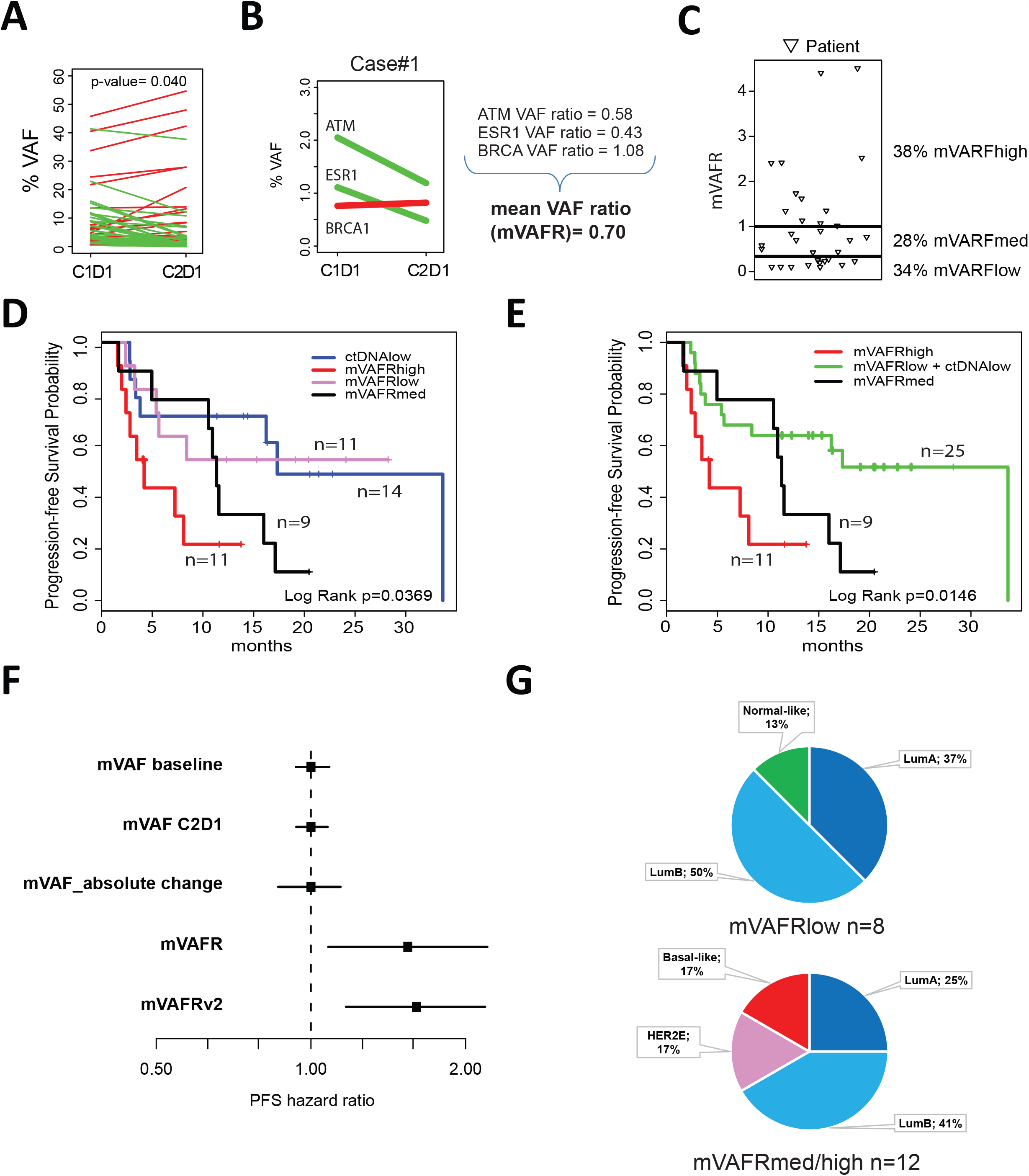
ctDNA dynamics and survival outcome. **(A)** VAF changes of 80 detected mutations from baseline to C2D1; **(B)** Illustration of a case and its mVAFR; **(C)** Distribution plot of patients with mVAFR-low (mVAFR of ≤0.3), mVAFR-medium (mVAFR of 0.31-0.99) and mVAFR-high (mVAFR of ≥1.0); **(D)** PFS based on ctDNA dynamics; **(E)** PFS based on ctDNA dynamics after combining the VAFR-low and ctDNA-low groups; **(F)** PFS hazard ratio forest plots across 5 different methods of assessing mVAF as a continuous variable: mVAF baseline, mVAF at C2D1, absolute change of mVAF between baseline and C2D1, mVAFR of all mutations with a VAF ≥0.4 at baseline and mVAFRv2 (mVAFR of all mutations including those with a VAF <0.4 at baseline); **(G)** Distribution of the intrinsic subtypes in the mVAFR-low group and the mVAFR-medium/high group (LumA=Luminal A, LumB=Luminal B, HER2E=HER2-Enriched).

With a median follow-up of 20.4 months, a significant association between the various ctDNA groups and PFS was observed across all patients (**Fig. 2D** and **2E**). Compared to the mVAFR-high group (i.e. low ctDNA response), the mVAFR-low group (i.e. high ctDNA response) was associated with better PFS (not reached vs. 4.2 months; hazard ratio [HR]=0.29, 95% confidence interval [CI] 0.1-0.9, p-value=0.034). Similarly, the mVAFR-low and ctDNA-low groups combined was associated with better PFS compared to the mVARF-high group (16.0 vs. 4.2 months; hazard ratio [HR]=0.26, 95% CI 0.1-0.7, p-value=0.007). The results in ctDNA-low group are in line with what was previously described in other metastatic tumors, as low ctDNA levels seem to be a good prognostic feature^17,18^. In addition, mVAFR as a continuous variable was also found significantly associated with PFS (HR per 1-unit increase=1.6, 95% CI 1.2-2.2, p-value=0.003). Of note, mVAF at baseline, or mVAF at C2D1 or absolute changes in mVAF were not found associated with PFS when evaluated as continuous variables (**Fig. 2F**). Interestingly, no patient with non-Luminal tumors (i.e. Basal-like or HER2-enriched [HER2-E]) was identified as high ctDNA responder (i.e. mVAFR-low). In patients with a medium or low ctDNA response (i.e. mVAFR-medium or mVAFR-high), 34% had non-Luminal tumors (i.e. HER2-E or Basal-like), consistent with previous reports in advanced HR+/HER2-negative disease associating the non-Luminal phenotype with poor prognosis^19,20^ and lack of response to ET^21,22^.

At baseline, median CA-15.3 value was 45 U/mL (7-6,672), and 26 patients (61%) had high CA-15.3 values (> 35 U/mL). At C2D1, median CA-15.3 value was 42 U/mL (range 8-9,868), and 19 patients (51%) had high CA-15.3 values. No significant differences in CA-15.3 levels were observed between baseline and C2D1 (p-value=0.350). The median ratio of CA-15.3 between C2D1 and baseline was 1.05 (range 0.4-1.6). No correlation was observed between CA-15.3 ratio and ctDNA mVFAR (correlation coefficient = −0.021). The levels of CA-15.3 at baseline or C2D1, and the CA-15.3 ratio, were not found associated with PFS (data not shown).

Our study has limitations worth noting. First, the limited sample size, which precludes more in-depth analysis within subgroups of patients. For example, identification of an optimal mVAFR cutoff to define prognosis will require a larger sample set. Second, the short follow-up time does not allow associations with overall survival. Third, we did not explore if tracking ctDNA levels of particular genes is better than tracking any detected altered gene in plasma. Nonetheless, a strong argument in favor of our approach is that it does not rely on specific genes but rather on the dynamic changes of the altered genes identified before initiating treatment. Fourth, it is unclear if this approach and methods will be applicable for other therapies or other cancer-types. Fifth, we cannot exclude that some of the alterations identified are from clonal hematopoyesis^23^. Finally, our findings will require further validation in retrospective or prospective cohorts of patients with advanced breast cancer treated with CDK4/6i and ET.

Our findings have several potential clinical implications. Most importantly, they suggest that early ctDNA dynamics using a multi-gene assay and a particular statistical methodology (i.e. mVAFR) serve as a general biomarker to identify patients with advanced breast cancer who are at high risk of progression during standard therapy with CDK4/6i and ET, giving the opportunity to intervene and change the treatment or add another treatment early. Notably, the biomarker seems independent of baseline clinical features, tumor marker CA-15.3 and the clinical setting (i.e. first line versus later lines), and the relationship of low ctDNA-responders with the non-luminal subtypes is weak. It will be important to also assess its value on the outcome of additional therapies and cancer types. Overall, our findings support the notion that monitoring ctDNA should be an integral part during drug development and should allow the design of novel clinical trials in key patient populations, such as those patients with an unfavorable ctDNA response.

## Methods

Detailed information on the experimental design can also be found in the Nature Research Reporting Summary.

All data generated or analyzed during this study are included in this published article (and its supplementary information files).

### Study design and patients

This is a prospective, single-center study in 50 consecutive patients with advanced breast cancer. Eligible patients were ≥18 years of age with histologically confirmed HR+/HER2-negative inoperable or metastatic breast cancer treated with a CDK4/6 inhibitor and ET. Blood samples for sample collection were obtained at baseline and at cycle 2 day 1 (C2D1). Clinical data, results of computed tomography (CT) imaging, and serial blood samples were collected as per standard practice. The study was performed in accordance with Good Clinical Practice guidelines and the World Medical Association Declaration of Helsinki. The study was approved by the local institutional research ethics committee, and all patients provided written informed consent.

### Plasma samples

Approximately 30 mL of venous blood was extracted at each time-point and collected in EDTA tubes. Blood was processed within 2 hours after the collection. Centrifugation at 1600g for 10 minutes at 4°C was performed to separate the plasma from the peripheral-blood cells. We obtained approximately 12 mL of plasma per patient and time-point, and plasma was immediately aliquoted in 1.5mL tubes and then we centrifugated them at 16000g at 4°C for other 10 minutes to remove the residual supernatant and any remaining contaminants including cells. Separated plasma was aliquoted in a 1.5 mL tube and immediately stored in a deep freezer at −80 °C.

Cell-free DNA (cfDNA) was extracted from 1.5 ml aliquots of plasma using the QIAamp circulating nucleic acid kit (Qiagen), concentrated using Agencourt Ampure XP beads (Beckman Coulter), and quantified by Qubit fluorometer (Life Technologies, Carlsbad, CA, USA). All cfDNA isolation and sequencing was performed at Guardant Health (Redwood City, CA, USA).

### DNA sequencing

Genomic alterations (mutations, insertions, deletions, fusions and amplifications) were detected from cfDNA extracted from plasma samples using a broad targeted NGS-based 74-gene panel (Guardant360), including coverage of the most prevalent tumor suppressor genes in human cancers. After isolation of cfDNA by hybrid capture, the assay was performed using molecular barcoding and proprietary bioinformatics algorithms with massively parallel sequencing on an Illumina Hi-Seq 2500 platform in a CLIA/CAP accredited laboratory (Guardant Health; Redwood City, CA, USA).

### ctDNA response definition

We filtered somatic mutations with variant allele frequency (VAF) ≥0.4% either at baseline (cycle 1 day 1 [C1D1]) or C2D1, based on 95%–100% limits of detection for this technology. Those patients with VAF <0.4% at both time-points were considered low-shedding tumors. We calculated the proportional change for all variants detected between the 2 time-points (VAF ratio [VAFR] = VAF_C2D1 / VAF_C1D1). For variants detected at 1 time-point but not the other, VAF was set to 0. We considered all undetected variants at C2D1, or variants with a VAF <0.4% at C2D1, to have a VAFR of 0.1. The reason is to prevent skewing of the average by variance introduced by quantitation variability below 0.4% VAF. We considered all new variants detected at C2D1 but not at baseline to have a VAFR of 10. The reason is to prevent skewing of the average by variance introduced by quantitation variability below 0.4% and by dividing by numbers that approach zero. Finally, a mVAFR was calculated per patient taking the average of all VAFR.

### CA-15.3 determination

The CA-15.3 assay was performed by the BRAHMS Kryptor Plus compact controller using TRACE (Time-Resolved Amplified Cryptate Emission) technology. CA-15.3 was considered elevated when it was above the normal upper limit (35 U/mL).

### PAM50 subtype determination

A minimum of ∼125 ng of total RNA from formalin-fixed paraffin embedded tumor samples was used to measure the expression of the 50 PAM50 subtype predictor genes and 5 housekeeping genes using the nCounter platform (Nanostring Technologies, Seattle, USA). Normalization of gene expression and PAM50 subtyping was performed as previously described^24^.

### Statistical analysis

The primary objective was to evaluate the association of ctDNA dynamics from baseline to C2D1 and PFS. PFS was defined as the time from initiation of treatment until progression or death. Univariate Cox proportional hazard regression analysis was used to investigate the association of each variable with PFS. The significance level was set to a 2-sided alpha of 0.05. All analyses were performed with R code 3.6.3.

## Data Availability

The data that support the findings of this study are available from the corresponding
author upon reasonable request.

## Contributors

AP had the idea for and designed the study. AP, OM, TP, FBM, NC, MM, MVL, BA, PG, DM, AR contributed to data collection and assembly. JIO, AP, OM and IF interpreted and analyzed data. All authors wrote and reviewed the report and approved the final version for submission.

## Acknowledgments

This study was funded by Instituto de Salud Carlos III (to A.P.), Breast Cancer Research Foundation (to A.P.), PhD4MD (to N.C.), Fundació La Marató TV3 (to A.P), RESCUER Horizon 2020 (to A.P.), Save the Mama (to A.P.), Pas a Pas (to A.P.), Asociación Cáncer de Mama Metastásico (to A.P.), Fundación Científica Asociación Española Contra el Cáncer (to F.B.M.) and Fundación SEOM (SEOM 2018 Grant: Fellowship for Training in Research in Reference Centers to T.P.).

